# Can Large Language Models Recognize Primary Immunodeficiency from Patient-Described Symptoms?

**DOI:** 10.64898/2026.05.26.26353818

**Authors:** Leon C. Reteig, Steven Woloshin, Paul J. Maglione, Jocelyn R. Farmer, Mei-Sing Ong

## Abstract

Patients with primary immunodeficiency (PI) face prolonged diagnostic delays and may increasingly turn to large language models (LLMs) to interpret their symptoms during this period. We evaluated whether an LLM could recognize PI from symptom descriptions derived from interviews with 21 PI patients. In prior work, we showed that GPT-4o identified PI in 96% of cases when prompted with physician-written patient histories (Rider *et al*., 2025). Here, when prompted with symptom descriptions in patients’ own words, GPT-5.2 identified PI in only 7 cases (33%), although it suggested general immune system concerns in 17 cases (81%). This expert-versus-patient gap may indicate that LLMs are sensitive to the language and framing of symptom descriptions, performing substantially worse when patients describe their own symptoms in everyday language than when clinicians summarize patient histories in structured medical terms. This study underscores the need to carefully evaluate how LLMs are used in patient-facing applications.

---

One in three US adults now uses large language model (LLM)-based chatbots for health information^1^, with ChatGPT alone fielding billions of such queries every week^2^. Symptom assessment accounts for 10%^3^ to 27%^1^ of these interactions. Patients with rare diseases are especially likely to turn to LLMs for guidance, as prolonged diagnostic odysseys and difficulty finding clinicians familiar with their conditions leave many searching for answers on their own.

Patients with primary immunodeficiencies (PI), also known as inborn errors of immunity, exemplify this scenario. PI is a collection of rare diseases with a prevalence of up to 1:25,000 individuals for the most common form^4^, characterized by recurrent infections, a heightened risk of autoimmune/autoinflammatory complications, and malignancy. Definitive diagnosis requires specialty immunological workup and exclusion of alternative causes, which is challenging for non-specialist physicians. As a result, many patients experience diagnostic delays of several years, which significantly impacts their quality of life and contributes to preventable morbidity.^5^ These patients are likely to seek LLM guidance while navigating their diagnostic odyssey. Whether LLMs can reliably support these patients, however, remains uncertain.

Emerging research has shown that LLM-generated responses to health-related queries often do not meet quality standards^6^, and that response quality can vary with subtle changes in prompts^7^. Published studies evaluating LLMs for disease diagnosis have almost exclusively relied on prompts crafted by medical experts, with limited attention to how these tools perform when used by members of the general public.^8^ Johri et al. showed that simulating patient-LLM interactions reduced diagnostic performance, compared to providing the LLM with direct clinical information.^9^ Likewise, Bean et al. showed that LLMs score much higher on emergency triage tasks when prompted with expert-written cases than when those cases are relayed by members of the general public.^7^ These findings suggest that LLMs may not be as performant in the hands of patients, although neither study directly examined actual patient use, instead relying on LLM-based patient simulations and healthy individuals as proxies.

We previously demonstrated that GPT-4o was able to recognize PI in 96% of patient cases when prompted with physician-written patient histories.^10^ Here, we assessed whether GPT-5.2 can recognize PI when prompted with symptom descriptions from patients.

This study used data collected from a qualitative study examining barriers to early diagnosis of PI. We conducted semi-structured qualitative interviews with adults (aged ≥18) diagnosed with PI who had experienced a delay in diagnosis. Participants were recruited from immunology clinics at two academic medical centers in Massachusetts (Beth Israel Lahey Hospital and Boston Medical Center) via leaflets distributed to PI patients by treating immunologists.

Recruitment followed a purposive sampling strategy with maximal variation for obtaining a heterogeneous sample with a range of age, sex, racial/ethnic group, medical history, and socioeconomic status. Given existing evidence that diagnostic delay is disproportionately experienced by underserved populations^11^, recruitment deliberately targeted those patients, including racial and ethnic minoritized groups, individuals with physical or intellectual disability, and those from low socioeconomic backgrounds. Recruitment continued until thematic saturation was reached, defined as the point at which additional interviews did not yield new themes relevant to the research questions. This study was approved by the Boston Children’s Hospital Institutional Review Board (IRB #P00050531).

For the current analysis, we extracted responses to a specific interview question: “What were your initial symptoms that led you to believe something was wrong?”. These symptom descriptions were then input into GPT-5.2. Patient responses were fully de-identified, and any mentions of their eventual PI diagnosis were redacted to ensure the LLM was evaluating symptoms alone without diagnostic information. We evaluated whether the LLM included PI in the differential diagnosis and whether it suggested broader immune system concerns. Table 1 presents representative examples of patient symptom descriptions, extracted verbatim from the interview transcripts. Detailed Methods are provided in the Supplementary Materials.

**Table 1.**
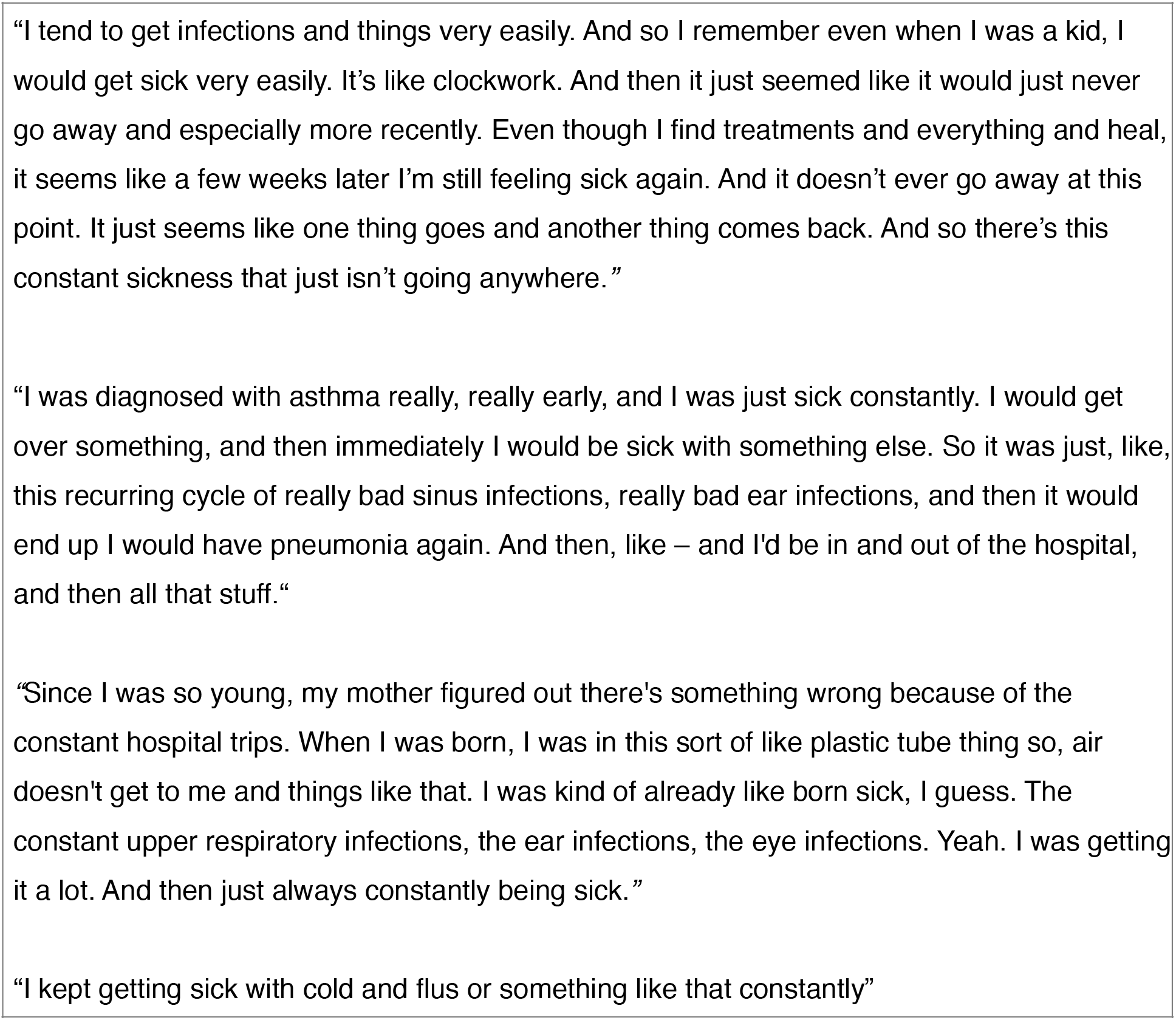
Representative examples of patient symptom descriptions, extracted verbatim from interview transcripts

The analysis included 21 patients with PI diagnoses confirmed by an immunologist (15 female, 6 male). Self-reported race was White (n=14), Black (n=3), Native American (n=2), Asian (n=1), and unknown (n=1); 2 patients identified as Hispanic. The most common diagnosis was common variable immunodeficiency (n=10/21, 48%). The other diagnoses were predominantly antibody deficiency (n=6), idiopathic CD4 lymphopenia (n=2), autoimmune polyendocrinopathy-candidiasis-ectodermal dystrophy (n=2), and combined immunodeficiency (n=1).

GPT-5.2 included PI in the differential diagnosis in 33% (7/21) of cases, substantially lower than the 96% accuracy achieved with physician-crafted prompts in our previous study^10^. More broadly, GPT-5.2 suggested a “weakened immune system” in 67% (14/21) of cases, and any potential “immune system issues” (without a specific diagnosis) in 81% (17/21) of cases. In all cases where PI was included in the differential diagnosis, patient descriptions included at least one of three classic features of PI (Table 2): (1) infections since childhood, (2) serious infections (e.g., sepsis), or (3) non-infectious manifestations (e.g., failure to thrive, gastrointestinal symptoms). The LLM generated an average of 14 differential diagnoses per patient. Beyond PI and general immune system concerns, the most frequent alternative diagnoses suggested by the LLM included: *allergic/atopic disease* (n=13; 62%; e.g. asthma and allergic rhinitis); *environmental/lifestyle factors* (n=13; 62%; e.g. daycare/school exposure, stress, sleep deprivation, poor air quality; *endocrine/metabolic disease* (n=9; 43%; e.g. diabetes, thyroid disorders, *nutritional deficiencies*); *autoimmune disease and chronic inflammatory conditions* (n=9; 43%); *anatomical/structural issues* (n=8; 38%; e.g. eustachian tube dysfunction, enlarged adenoids, deviated septum); and *genetic/congenital syndromes* (n=7; 33%; *e*.*g*. cystic fibrosis, primary ciliary dyskinesia). These categories closely reflect the differential diagnoses that are routinely considered in immunology clinics when evaluating for PI.

**Table 2.** Patient symptom descriptions and LLM-generated differential diagnoses

| Symptom Pattern | n (%) | LLM-generated differential diagnosis |  |
| --- | --- | --- | --- |
|  |  | Mentions PI specifically, n (%) | Mentions general immune system issues without specific diagnosis, n (%) |
| Recurrent infections (no childhood onset specified) | 8/21 (38%) | 2/8 (25%) | 5/8 (62%) |
| Recurrent infections since childhood | 9/21 (43%) | 4/9 (44%) | 4/9 (44%) |
| Serious/severe infections<br>(Required hospitalizations, IV antibiotics, sepsis, bacteremia, pneumonia) | 5/21 (24%) | 3/5 (60%) | 1/5 (20%) |
| Non-infectious manifestations only | 3/21 (14%) | 1/3 (33%) | 1/3 (33%) |
| **TOTAL** | 21/21 (100%) | 7/21 (33%) | 17/21 (81%) |

**Note:** Patients with a mention of primary immunodeficiency (PI) are not included in the “immune system issues” column. In contrast, symptom patterns are not mutually exclusive; patients may appear in multiple rows.

This study reveals a substantial gap between LLM diagnostic performance when prompted by medical experts (96%) versus patients (33%), highlighting that the promise of LLMs for diagnosis may depend heavily on how symptoms are communicated. However, the broader category of “immune system issues” (81%) may still provide value, especially since these patients experienced prolonged diagnostic delay and standard clinical evaluation initially missed the diagnosis. While not diagnostically definitive, the suggestion of “immune system issues” could help patients advocate for specialist evaluation or more thorough workup.

Our findings are consistent with a recent study demonstrating that while LLMs performed well on medical knowledge benchmarks when tested in isolation (94.9% accuracy), performance dropped dramatically when these models were used by members of the public (34.5% accuracy).^7^ As LLMs become an increasingly routine source of medical information for patients and caregivers, this gap represents not just a missed opportunity, but a patient safety risk.

The study has several limitations. First, patients were interviewed post-diagnosis, raising concerns about recall bias and whether descriptions reflect what was reported during initial clinical encounters. Participants aware of their diagnosis may selectively recall immune-related symptoms, providing more diagnostically informative descriptions than they would have pre-diagnosis. Our findings therefore likely represent a best-case estimate; LLM performance in the intended use case among patients seeking guidance before diagnosis may be lower. Second, the small sample size of 21 patients, though large for a rare condition, does not capture the full spectrum of PI presentations. Furthermore, cohort selection based on documented delayed diagnosis may enrich the cohort for presentations that challenged both clinicians and LLMs. The LLM appeared to identify PI primarily when patients explicitly mentioned classic features of PI, suggesting potential difficulty recognizing more subtle presentations, atypical symptom patterns, adult-onset PI, or autoimmune/inflammatory manifestations. Most critically, while the study evaluated patients with confirmed PI, we did not assess specificity by testing patient descriptions from those without PI. The clinical utility of flagging “immune system issues” depends critically on whether this occurs at unacceptably high rates in healthy individuals or those with other conditions. PI is a rare disease and there is already a significant shortage of immunologists. Generating excessive false alarms could further strain limited specialist resources and delay care for patients who truly need evaluation.

Nonetheless, our study addresses a critical gap in the LLM diagnostics literature by using actual patient language rather than expert-crafted prompts, better reflecting how patients might interact with LLMs in practice. An ecologically valid evaluation of LLM diagnostic performance is inherently challenging, requiring prospective capture of LLM queries from large numbers of patients with undiagnosed conditions and longitudinal follow-up to confirm their eventual diagnoses. Perhaps for this reason, most published evaluations of LLM diagnostic accuracy rely on standardized clinical vignettes (e.g., USMLE-style cases) or expert-crafted scenarios, which do not reflect how patients describe their symptoms. Our approach using retrospective interview data represents a pragmatic middle ground, capturing more authentic patient language than expert prompts, while remaining feasible to conduct.

Our findings underscore the need to evaluate LLMs under realistic patient-use conditions before promoting them as tools to support diagnosis of rare diseases. As adoption of LLMs by the general public outpaces evidence, further research is needed to characterize the consequences and unintended harms of patients turning to LLMs to guide their diagnostic journey.

“I tend to get infections and things very easily. And so I remember even when I was a kid, I would get sick very easily. It’s like clockwork. And then it just seemed like it would just never go away and especially more recently. Even though I find treatments and everything and heal, it seems like a few weeks later I’m still feeling sick again. And it doesn’t ever go away at this point. It just seems like one thing goes and another thing comes back. And so there’s this constant sickness that just isn’t going anywhere.*”* “I was diagnosed with asthma really, really early, and I was just sick constantly. I would get over something, and then immediately I would be sick with something else. So it was just, like, this recurring cycle of really bad sinus infections, really bad ear infections, and then it would end up I would have pneumonia again. And then, like – and I’d be in and out of the hospital, and then all that stuff.” *“*Since I was so young, my mother figured out there’s something wrong because of the constant hospital trips. When I was born, I was in this sort of like plastic tube thing so, air doesn’t get to me and things like that. I was kind of already like born sick, I guess. The constant upper respiratory infections, the ear infections, the eye infections. Yeah. I was getting it a lot. And then just always constantly being sick.*”* “I kept getting sick with cold and flus or something like that constantly”

## Data Availability

The data supporting the findings of this study consist of patient interview transcripts and are available from the corresponding author upon reasonable request, subject to institutional data sharing agreements and ethical approval. The codebook used for annotation is provided in the supplementary material.

## Acknowledgments

Research reported in this publication was supported by funding from the National Institute on Minority Health and Health Disparities of the National Institute of Health under award number R01MD017816. The content is solely the responsibility of the authors and does not necessarily represent the official views of the National Institutes of Health. We are grateful to the patients who participated in our interview study and generously shared their experiences.

## Notes

### Competing Interest Statement

PJM has served as a consultant for ADMA Biologics, Novartis, Pharming, and Sanofi, and has received investigator-initiated research funding from Pharming and Takeda. JRF has served as a consultant for Pharming, has received investigator-initiated research funding from Bristol Myers Squibb, Pfizer, and Pharming, and serves as an Associate Editor for a Springer Nature publication. LCR and MSO have received investigator-initiated research funding from Pharming, unrelated to the present work. SW serves on the editorial board of JAMA Internal Medicine and on the board of Preventing Overdiagnosis, both in an unpaid capacity.

### Author Declarations

IRB of Boston Children's Hospital waived ethical approval for this work

### Summary of Updates

Article rewritten for non-specialist (immunologist) audience

