## Supplementary Methods for "Can Large Language Models Recognize Primary Immunodeficiency from Patient-Described Symptoms?"

### Supplementary information

#### Interview data

This study used interview data from a prior qualitative study aimed at understanding barriers to early diagnosis of primary immunodeficiency (PI). Semi-structured interviews were conducted by telephone using an interview guide informed by the Model of Pathway to Treatment.[1] Participants were asked to describe their diagnostic journey and the challenges they faced in obtaining a timely diagnosis of PI. Interviews were audiotaped and transcribed verbatim. All participants received a $100 gift card upon completion. A total of 22 patients were recruited; 21 interviews were conducted in English and one in Spanish. We excluded the Spanish-language interview from this analysis, as translation could alter the lay language patterns central to our research question.

#### LLM querying

For the current study, we extracted patients' responses to the following question: "What were your initial symptoms that led you to believe something was wrong?" We removed all identifiers and any mention of patients' eventual PI diagnosis to ensure the LLM was only given information about initial presentation. The resulting text snippets had a median word count of 42 (IQR: 57) and contained an average of 3.1 sign/symptom descriptions per patient. These texts were input without further modification into a private deployment of GPT-5.2 using the Azure OpenAI Service (queries conducted in

April 2026). The Azure OpenAI Service operates under a data processing agreement that prevents submitted data from being used to train or improve foundation models and ensures data is not shared with OpenAI or other third parties. Although all inputs had been de-identified prior to submission, this private deployment provided an additional safeguard for patient data confidentiality.

**Annotation and analysis**

One author (LCR) annotated all signs/symptoms in the inputs and all differential diagnoses in the LLM output using Taguette (version 1.5.2).[2] The annotations were reviewed by a second author (M-SO) and discrepancies were resolved through discussion until consensus was reached. The full codebook including all annotated symptom and diagnosis categories with frequencies is provided in Table E1. The resulting annotations were analyzed using the R programming language (version 4.4.3; 2) and the *here* (version 1.0.1; 2), *janitor (version 2.2.1; 3)* and *tidyverse* (version 2.0.0, 4) packages.
